# The copy number variant architecture of psychopathology and cognitive development in the ABCD^®^ study

**DOI:** 10.1101/2024.05.14.24307376

**Authors:** Zhiqiang Sha, Kevin Y. Sun, Benjamin Jung, Ran Barzilay, Tyler M. Moore, Laura Almasy, Jennifer K. Forsyth, Smrithi Prem, Michael J. Gandal, Jakob Seidlitz, Joseph T. Glessner, Aaron F. Alexander-Bloch

## Abstract

**Importance:** Childhood is a crucial developmental phase for mental health and cognitive function, both of which are commonly affected in patients with psychiatric disorders. This neurodevelopmental trajectory is shaped by a complex interplay of genetic and environmental factors. While common genetic variants account for a large proportion of inherited genetic risk, rare genetic variations, particularly copy number variants (CNVs), play a significant role in the genetic architecture of neurodevelopmental disorders. Despite their importance, the relevance of CNVs to child psychopathology and cognitive function in the general population remains underexplored.

**Objective:** Investigating CNV associations with dimensions of child psychopathology and cognitive functions.

**Design, Setting, and Participants:** ABCD**^®^** study focuses on a cohort of over 11,875 youth aged 9 to 10, recruited from 21 sites in the US, aiming to investigate the role of various factors, including brain, environment, and genetic factors, in the etiology of mental and physical health from middle childhood through early adulthood. Data analysis occurred from April 2023 to April 2024.

**Main Outcomes and Measures:** In this study, we utilized PennCNV and QuantiSNP algorithms to identify duplications and deletions larger than 50Kb across a cohort of 11,088 individuals from the Adolescent Brain Cognitive Development**^®^** study. CNVs meeting quality control standards were subjected to a genome-wide association scan to identify regions associated with quantitative measures of broad psychiatric symptom domains and cognitive outcomes. Additionally, a CNV risk score, reflecting the aggregated burden of genetic intolerance to inactivation and dosage sensitivity, was calculated to assess its impact on variability in overall and dimensional child psychiatric and cognitive phenotypes.

**Results:** In a final sample of 8,564 individuals (mean age=9.9 years, 4,532 males) passing quality control, we identified 4,111 individuals carrying 5,760 autosomal CNVs. Our results revealed significant associations between specific CNVs and our phenotypes of interest, psychopathology and cognitive function. For instance, a duplication at 10q26.3 was associated with overall psychopathology, and somatic complaints in particular. Additionally, deletions at 1q12.1, along with duplications at 14q11.2 and 10q26.3, were linked to overall cognitive function, with particular contributions from fluid intelligence (14q11.2), working memory (10q26.3), and reading ability (14q11.2). Moreover, individuals carrying CNVs previously associated with neurodevelopmental disorders exhibited greater impairment in social functioning and cognitive performance across multiple domains, in particular working memory. Notably, a higher deletion CNV risk score was significantly correlated with increased overall psychopathology (especially in dimensions of social functioning, thought disorder, and attention) as well as cognitive impairment across various domains.

**Conclusions and Relevance:** In summary, our findings shed light on the contributions of CNVs to interindividual variability in complex traits related to neurocognitive development and child psychopathology.

**Key Points:** *Question:* Are copy number variants (CNVs) contributing to individualized variability in psychopathology outcomes and cognitive functions in children?

*Findings:* Both regional CNVs at 1q12.1, 10q26.3 and 14q11.2 and global CNV burden accounted for the variance in dimensions of psychopathology, particularly for attention and social problems, as well as cognitive performance, especially for working memory and reading ability.

*Meaning:* Our findings suggest that, in addition to common genetic variants, gene dosage changes confer genetic susceptibility to child psychopathology and cognitive development.

## Introduction

Neurodevelopmental disorders are characterized by a spectrum of mental health issues, developmental delays, and deficits in diverse cognitive functions^1,2^. Genetic factors are known to play a crucial role in these disorders^3,4^ such as autism spectrum disorder^5,6^, attention-deficit/hyperactivity disorder^7,8^ and psychosis spectrum disorders^9^. Population-based genetic studies have revealed robust associations between common variants, such as single-nucleotide polymorphisms (SNPs), and the pathogenesis of these conditions^10,11^. The advent of genome-wide association scans has shed light on the polygenic nature of neurodevelopmental disorders. However, the contribution of SNPs with subtle genetic effects does not fully account for the variance in these complex phenotypes due to genetic factors ^12^. In addition to common genetic variants, other types of genetic variations including copy number variants (CNVs) – a form of genomic structural rearrangement involving duplications and deletions spanning at least 1000 base pairs throughout the human genome^13^ – are increasingly recognized as crucial components in elucidating the detailed genetic architecture underlying the etiology of neurodevelopmental disorders^14^.

While the large majority of CNVs are benign, clinical and genetic epidemiological investigations have consistently demonstrated that individuals harboring pathogenic CNVs often exhibit early-onset symptoms of mental illness and are more predisposed to transitioning into clinically diagnosable psychiatric disorders^15^. For example, approximately one quarter of individuals with the 22q11.2 deletion develop schizophrenia^16,17^. Large-scale case-control studies have identified a spectrum of genomic duplications and deletions associated with increased genetic susceptibility to neurodevelopmental disorders, including schizophrenia, attention-deficit/hyperactivity disorder, and autism spectrum disorder^5,6,18^. However, these investigations have predominantly focused on identifying genetic correlates within the confines of a categorical diagnostic framework, while it is increasingly acknowledged that psychopathological manifestations and associated cognitive deficits exist along a continuum beyond traditional diagnostic boundaries^19–21^. To date, there remains a dearth of research exploring the CNV architecture underlying dimensions of psychopathology and cognitive functions during childhood.

The concept of the CNV risk score has emerged to estimate the tolerance to loss-of-function mutations and dosage sensitivity of genes by deletions or duplications^22^. CNV risk scores aim to measure the cumulative risk from deletions or duplications of large genomic segments. Different types of genetic variants exhibit cumulative effects, contributing to overall phenotypic variability^12^. Recently, we observed that higher CNV risk scores were significantly correlated with poor cognitive performance and greater psychopathology in youth^23^. These findings underscore the potential of CNV risk scores to detect genetic associations with neurodevelopmental outcomes in childhood and adolescence.

In the present study, we conducted a comprehensive exploration of the CNV architecture underlying dimensions of psychopathology and cognitive phenotypes within the Adolescent Brain Cognitive Development (ABCD**^®^**) Study. This prospective study comprises a community sample of N=11,876 children aged 9-10 years old, who were not ascertained for mental health problems^24^. Our investigation proceeded in several stages. First, CNVs for each individual were identified using two algorithms, PennCNV^25^ and QuantiSNP^26^, leveraging genome-wide SNP array data. Second, CNVs that overlapped between the two algorithms were utilized in a genome-wide CNV association scan, aiming to identify specific CNV regions (CNVRs) associated with psychopathology and cognition independently. Third, we examined whether individuals carrying CNVs associated with neurodevelopmental disorders^22^ exhibited poorer cognitive performance and greater psychopathology. Finally, we explored the association of CNV risk score with subdomains of psychopathology and cognition. Cumulatively, the present study sheds significant light on the contribution of CNVs to interindividual variability in complex traits critical to neurodevelopmental outcomes.

## Methods

### Sample and CNV quality control

We downloaded the imputed genotyped data from the ABCD**^®^** portal (Data Release 3.0). These genetic data were acquired by the Chemagen bead-based/Chemagic STAR DNA Saliva4k Kit (CMG-1755-A) at the Rutgers University Cell and DNA Repository^27^. Genotypes were ascertained using the Affymetrix National Institute on Drug Abuse SmokeScreen Array^28^, and imputed using SHAPEIT3^29^ and impute2^30^.

To call CNVs using ABCD**^®^**study, we compiled log R ratio and B allele frequency profiles for each SNP marker from the intensity files of genotyping calls obtained through the Affymetrix Smokescreen array^27^. Multiple sample log R ratio and B allele frequency matrix files, paired with sample and probe information files were split into single sample log R ratio and B allele frequency files. Population frequency of B allele and GC base content model definition files specific to the Affymetrix SmokeScreen Array were compiled. CNV calling was performed following established procedures detailed in our previous publications (Supplementary Figure 1)^23,31^. The ABCD**^®^** study encompasses genotype data from 138 plates. Initially, individuals from plate 461 were excluded due to potential sequencing issues identified in the genetic data report. Additionally, individuals were excluded if their data exhibited a standard deviation of log R ratio >0.35, a standard deviation of B allele frequency >0.1, or an absolute value of wave factor >0.05. All probe coordinates were established based on the GRCh37/hg19 reference genome.

We employed hidden Markov models to call CNVs based on SNP genotype data, using both PennCNV^25^ and QuantiSNP^26^ algorithms. PennCNV utilizes multiple sources of information, including the log R ratio and B allele frequency at each SNP marker, the distance between neighboring SNPs, and adjusts for “genomic waves” using a regression model based on GC content^25^. On the other hand, QuantiSNP utilizes an objective Bayes approach, leveraging the log R ratio and B allele frequency for each SNP marker^26^. Both algorithms have demonstrated sensitivity in detecting deletions and duplications in the human genome across genetic studies spanning the past two decades^9,32,33^. To ensure identifying reliable CNVs, we used CNVision^34^ to merge CNVs detected by two algorithms. Furthermore, we excluded individuals carrying more than 500 merged CNVs, as this threshold may indicate batch effects, genotyping errors, or extreme chromosome abnormalities. CNVs exhibiting >70% overlap between the two algorithms were retained for follow-up genetic analysis.

Subsequently, we implemented a series of additional filtering steps aimed at minimizing false discoveries. Specifically, we excluded CNVs smaller than 50,000 base pairs in size and filtered out CNVs spanning <20 SNPs or with PennCNV or QuantiSNP confidence scores <30^23^. CNVs demonstrating over 50% reciprocal overlap with segmental duplications, centromeric regions, telomeric regions, or the major histocompatibility complex region were excluded due to the complex genomic structure in these regions. CNVs exhibiting an equivocal number of copies between the two algorithms were also removed. Moreover, in families with multiple members, only one random family member was included in the subsequent analysis. The final sample included N=8,564 individuals (mean age=9.9 years, 4,532 boys)

### Psychopathological and cognitive measures

The ABCD**^®^** study includes comprehensive assessments of psychopathological and cognitive measures^35,36^. We used derived measures of dimensional psychopathology from the 119-item Child Behavioral Checklist^37^ completed for each individual. Parents rated the extent to which specific behaviors (e.g., “Destroys others’ things”) were characteristic of their child over the past 6 months, using a 3-point scale: 0 (“not true”), 1 (“somewhat or sometimes true”), or 2 (“very true or often true”). These 119 items are aggregated to generate raw composite scores for 8 scales: anxious/depressed, withdrawn/depressed, somatic complaints, social problems, thought problems, attention problems, rule-breaking behavior, and aggressive behavior. The sum of scores on these scales provided an overall measure of psychopathology burden, encompassing various behavioral syndromes.

Cognitive performance was assessed by the NIH Toolbox Neurocognitive Battery. The NIH Toolbox encompasses 8 domains^38,39^, including: fluid intelligence (reflecting the ability to solve abstract reasoning problems); dimensional change card sort (measuring ability to plan, organize, and execute goal-directed behaviors); flanker inhibitory control and attention (reflecting the ability to handle multiple environmental stimuli); list sorting working memory (quantifying the ability to store, manipulate, and hold new information); oral reading recognition test (capturing reading ability and academic achievement); picture vocabulary test (characterizing language and verbal intellect); picture sequence memory (representing episodic memory including the acquisition, storage, and retrieval of information); and pattern comparison processing speed (indicating the ability to process new information within a certain amount of time). Scores across these domains were aggregated to generate a composite score reflecting overall cognitive performance.

For each cognitive and psychopathological measure, we regressed the effects of age, sex, and batch by linear regression models. Subsequently, rank-based inverse normalization was applied to ensure the residuals following a normal distribution.

### Genome-wide CNVR association analysis

We utilized ParseCNV2^40^ to conduct genome-wide CNVR association analyses for each cognitive and psychopathological measure independently. This method involves mapping individual-level CNV calls to population-level probe-based CNV statistics (separately for deletions and duplications). Subsequently, the significance of each SNP marker was assessed using Fisher’s exact test and incorporating covariates using linear regression for quantitative (continuous) trait phenotypes. The resulting association statistics were then used to combine SNPs in proximity (default distance of 1MB) with comparable p-values (default threshold of 1 power of 10) into genomic regions referred to as “CNVRs”. This approach offers several advantages over methods that investigate genetic correlates for individual CNVs, increasing power and preventing redundant reporting of overlapping genomic regions associated with the phenotype. Previous studies have successfully applied this approach to identify genomic structural changes associated with brain disorders and physical health^41–43^.

### Multiple testing correction of significance

In contrast to genome-wide association scans involving SNPs, a p-value threshold of 5×10^−4^ was recommended to define genome-wide significance for CNV (spanning many nucleotides) association testing after surviving multiple testing corrections. We applied a Bonferroni-corrected threshold of p<2.5×10^−4^ (i.e., 5×10^−4^/2) to identify CNVRs contributing to the overall psychopathological and cognitive measures. For the 8 dimensions of cognitive measures, after accounting for correlations between these traits using matrix spectral decomposition, the effective number of traits was estimated to be 6. Consequently, we applied a significance level of p<8.3×10^−5^ (i.e., 5×10^−4^/6) to correct for multiple testing. Similarly, for the 8 dimensions of psychopathology, we set a p-value threshold of p<1×10^−4^ (i.e., 5×10^−4^/5).

### CNV annotation and CNV risk scores

The CNV risk score is used to estimate the pathogenic functional consequences of CNV deletions and duplications in individuals -- encompassing genetic burden, intolerance, and dosage sensitivity. CNV burden quantifies the number of genes covered by CNVs or the size of CNVs carried by an individual. Genetic intolerance refers to the annotations of genes within CNVs concerning their ability to withstand loss-of-function mutations. Dosage sensitivity characterizes the sensitivity to the deletion and duplication of CNVs.

We annotated each CNV based on two public databases^44,45^, scoring them in terms of genetic burden, intolerance, and dosage sensitivity (Supplementary Figure 2). Genetic burden was quantified by the size of the CNV and the number of overlapping genes within the CNV. Intolerance metrics included the probability of loss intolerance (pLI)^46^ and the loss of function observed/expected upper bound fraction (LOEUF). Specifically, pLI represents the likelihood of a gene being intolerant to loss-of-function mutations, ranging from 0 to 1. A higher pLI score indicates less tolerance to inactivation, implying potentially deleterious functional outcomes. LOEUF compares the observed and expected number of loss-of-function mutations for a gene in a reference population, with values ranging from 0 to 2. Genes with a LOEUF value less than 0.35 are typically considered intolerant. For CNVs, the cumulative inverse LOEUF (iLOEUF) of all encompassed genes is computed. Dosage sensitivity is measured by the probability of haploinsufficiency (pHI) and the probability of triplosensitivity (pTS). These metrics reflect the likelihood of a gene being sensitive to copy number gain or loss, respectively. Thus, the pHI of a deletion is computed as the sum of the pHI values of all genes overlapping with the deletion, while the pTS of a duplication is estimated by the sum of the pTS values of all genes within the duplication. At the individual level, each component of the CNV risk score is calculated as the sum of all deletions and duplications, respectively.

We used a linear regression model to investigate the association of each CNV risk score with individual cognitive and psychopathological measures across 8564 individuals (the size of the unrelated sample after quality control), while controlling for the effects of age, sex, and array batch. Prior to modeling, rank-based inverse normalization was conducted on each behavioral measure to ensure adherence to a normal distribution. To correct for multiple testing, false discovery rate (FDR) adjustment at a significance level of p<0.05 was applied separately for cognitive measures and psychopathological measures.

## Results

### The CNV landscape of the ABCD^®^ study

Following conservative quality control (Supplementary Figure 1), a total of 5760 CNVs were identified across 4111 individuals (Figure 1) while 4453 individuals did not carry any CNVs. Among the detected CNVs, 1391 (24.15%) were deletions (43 homozygous), while 4369 (75.85%) were duplications (161 with copy number >3). While 28.8% of individuals with a CNV carried a single deletion, instances of up to three deletions were observed in nine individuals. Similarly, 61.5% of individuals had one duplication (totaling 2527 out of 4111), with up to four duplications found in 18 individuals (Supplementary Figure 3). Summary statistics at the CNV level were as follows (Supplementary Figure 3): number of SNPs encompassed by deletions, mean=78 (range 20-1811); number of SNPs encompassed by duplications, mean=151 (20-12,635); deletion length, mean=300.99Kb (50Kb-6.14Mb); duplication length, mean=357.76Kb (50Kb-11.10Mb); number of genes within deletions, mean=1 (0-46); number of genes within duplication, mean=2 (0-51) .

**Figure 1.**
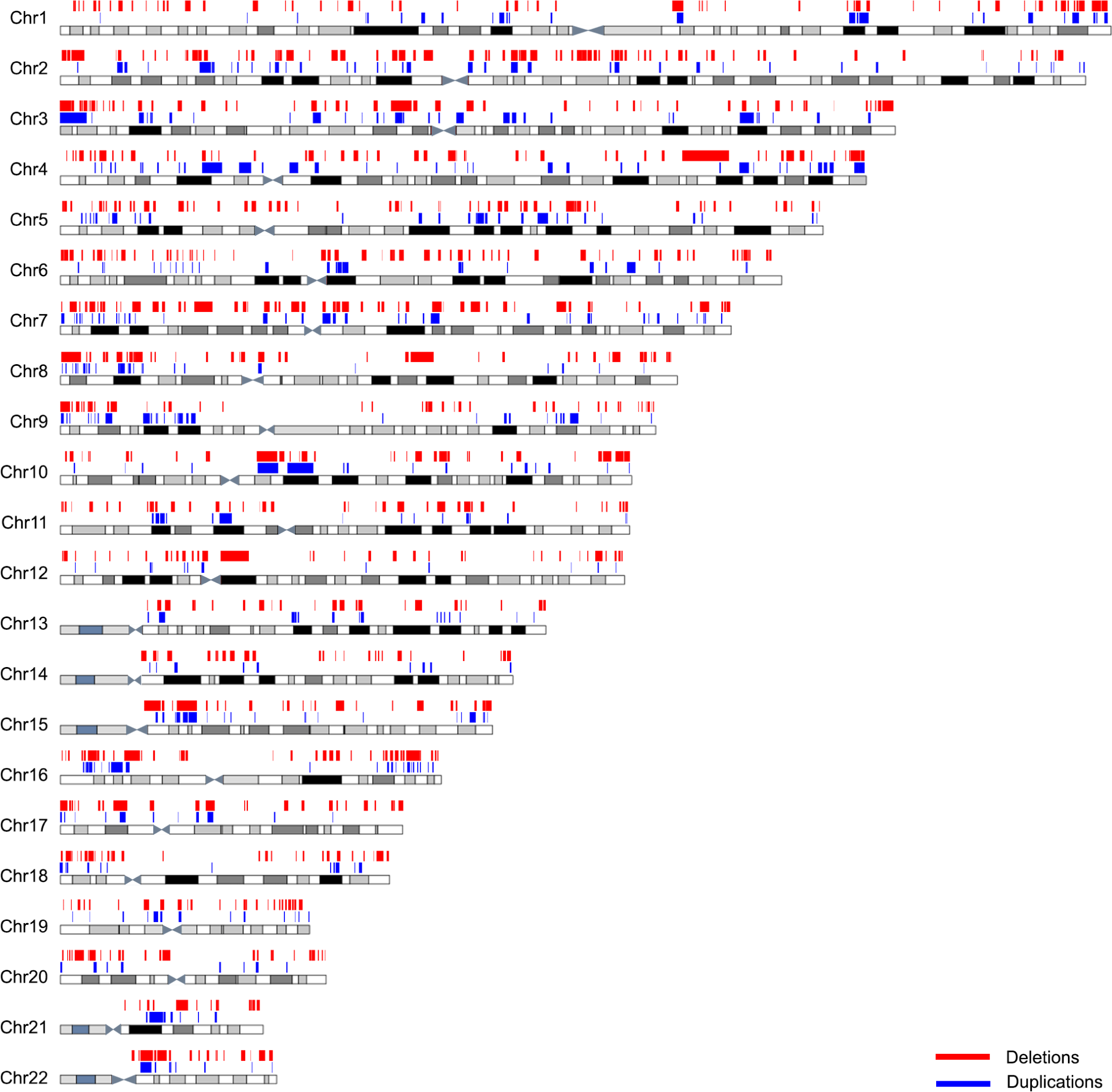
The landscape of the CNV calls in the ABCD^®^ study. After conservative quality control, we report 5760 CNVs among 8564 individuals from the ABCD**^®^** study. The figure illustrates the chromosomal distribution of all deletions and duplications called in the ABCD**^®^** study, in which blue represents deletions, and red represents duplications.

### Genome-wide CNVR association with overall psychopathology and cognition

We used ParseCNV2^40^ to identify CNVRs associated with variations in overall psychopathology and cognitive function, including one CNVR associated with overall psychopathology (Figure 2A) and three CNVRs associated with cognition (Figure 2B). Specifically, a duplication at 10q26.3 (i.e., chr10:133248899-133784881) was significantly associated with overall psychopathology (p=1.45×10^−4^, Figure 2A and Supplementary Table 1). Upon CNV decomposition, we identified 16 individuals carrying duplications that contributed to this significant CNVR association signal. Notably, two genes overlap with this genomic region, including *PPP2R2D* and the long non-coding RNA *AL450307.1 (*Figure 2A). Annotation revealed that common variants within *PPP2R2D* have been linked to schizophrenia^47^, and methylation modifications of the genomic region have been implicated in dimensional psychopathology in youth^48^. In a GWAS of 12 psychiatric disorders published by Romero et al.^49^, the strongest association signal was for rs10030847 (p=0.0003, Figure 2A).

**Figure 2.**
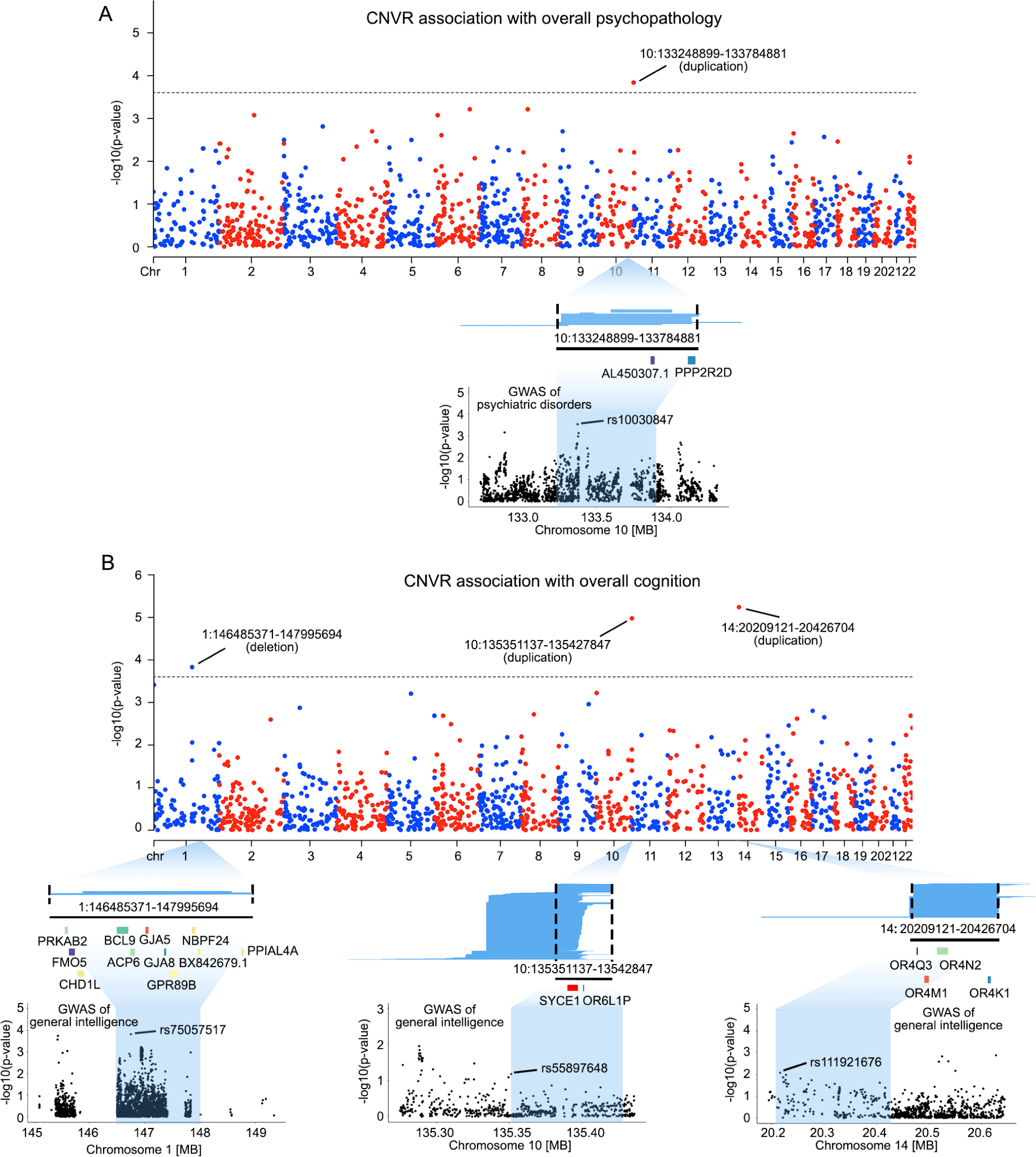
Genome-wide CNVRs associations with overall psychopathology and cognitive function. (A) Genome-wide CNVR association with overall psychopathology. Manhattan plot illustrates significant CNVR associations with the overall psychopathology measure at Bonferroni-corrected p<2.5×10^−4^ (i.e., 5×10^−4^/2), the recommended threshold for genome-wide significance in the ParseCNV2 platform. Blue lines under the Manhattan plot represent the 16 duplications contributing to the significant CNVR association. The black line and the genes below them indicate the genomic position of genes encompassed by the CNVR significantly associated with psychopathology. The Manhattan plot at the bottom illustrates the GWAS association of psychiatric disorders with common genetic variants (i.e., SNPs) within the identified CNVR, as reported by Romero et al^49^. (B) Genome-wide CNVR association with global cognitive function. Three CNVRs were found significantly associated with the overall cognitive function at Bonferroni-corrected p<2.5×10^−4^ (i.e., 5×10^−4^/2). Blue lines under the Manhattan plot represent the CNVs that contribute to each significant CNVR association. The black lines and the genes below them indicate the genomic position of genes encompassed by the CNVR significantly associated with global cognitive function. The Manhattan plot at the bottom illustrates GWAS SNP associations with general intelligence within each CNVR, as reported by Savage et al.^50^.

Three CNVRs were significantly associated with general cognition, including chr1:146485371-147995694 deletion at 1q21.1 (p=1.48×10^−4^), chr10:135351137-135427847 duplication at 10q26.3 (p=1.05×10^−5^), and 14:20209121-20426704 duplication at 14q11.2 (p=5.75×10^−6^, Figure 2B and Supplementary Table 2).

For the deletion at chr1:146485371-147995694, we identified 2 individuals carrying duplications that were associated with this CNVR. Notably, both individuals exhibited poor performance across multiple cognitive domains, particularly in cognitive flexibility, working memory, and language-related functions (Supplementary Figure 4). Regional log R ratio and B allele frequency plots confirmed that copy number loss was present at 1q22.1 in both individuals (Supplementary Figure 5). This CNVR encompasses several genes important for cognitive development, such as *PRKAB2, GJA8* and *CHD1L*^51–53^ (Figure 2B). Of note, 1q21.1 is a known pathogenic CNV associated with neurodevelopmental disorders and intellectual disability ^22,54,55^, providing a validation of the methodological approach. In a GWAS of general intelligence^50^, the top SNP in this CNVR was rs75057517 (p=0.0001, Figure 2B), indicating that common variants in the CNVR may also impact general intelligence.

For the duplication at chr10:135351137-135427847, we identified 717 individuals carrying copy number gains associate with this CNVR (Figure 2B). This region encompasses two protein-coding genes, *SYCE1* and *OR6L1P*. Notably, duplications in this CNVR have been previously reported to be a contributing factor for intellectual disability^56^. The top SNP in the CNVR in the general intelligence GWAS was rs55897648 (p=0.06, Figure 2B).

For the association of duplication at 14:20209121-20426704, we observed 350 individuals with duplications associated with this CNVR located at 14q11.2. This genomic region contains four olfactory receptor family genes, including *OR4Q3, OR4N2, OR4M1* and *OR4K1* (Figure 2B). Variations in the copy number of olfactory genes have been identified as significantly associated with cognitive decline in patients with Alzheimer’s disease^57^. The top SNP in the CNVR in the general intelligence GWAS was rs111921676 (p=0.008, Figure 2B).

### Genome-wide CNVR association with dimensions of psychopathology and cognition

Across 8 dimensions of psychopathology, we found that the 14q11.2dup CNVR was significantly associated with attention problems (p=6.39×10^−5^, Figure 3A, Supplementary Figure 6 and Supplementary Tables 3-10) and also had a marginal association with somatic complaints (p=1.45×10^−4^, Figure 3A and Supplementary Table 5). Across 8 domains of cognitive function, the 14q11.2dup CNVR was also significantly associated with fluid intelligence (p=3.91×10^−5^, Figure 3B and Supplementary Table 11) and reading (p=4.24×10^−5^, Figure 3C and Supplementary Table 15). In addition, the CNVR at chr10:135351137-135427847 was significantly associated with working memory (p=3.21×10^−5^, Figure 3D and Supplementary Table 13). For further details, see Supplementary Figure 7 and Supplementary Tables 11-18.

**Figure 3.**
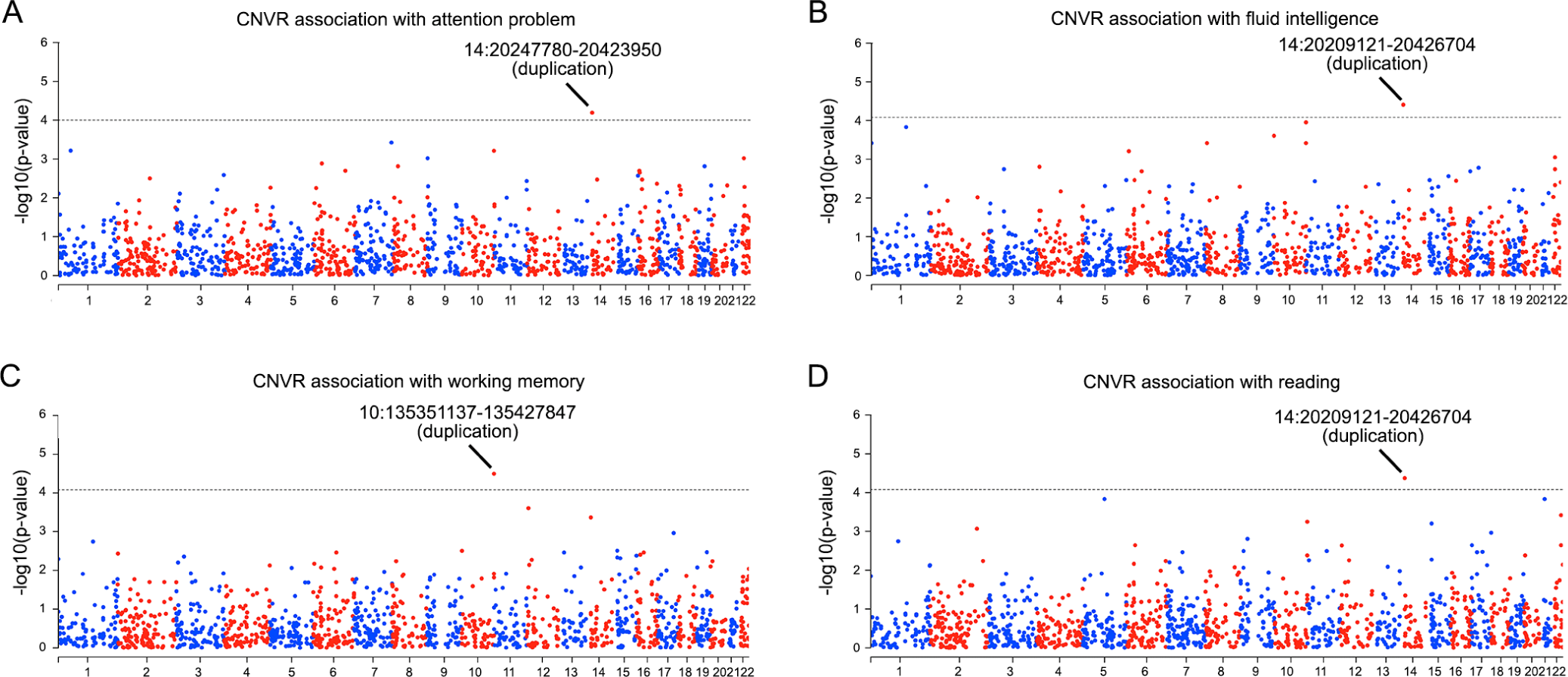
Genome-wide CNVRs associations with dimensions of psychopathology and cognitive function. A) Across 8 dimensions of psychopathology, we found duplication of 14:20247780-20423950 at 14q11.2 was significantly associated with attention problems at Bonferroni-corrected p<1×10^−4^ (i.e., 5×10^−4^ divided by 5, which is the effective number of tests across 8 dimensions of psychopathology). B-D) Across 8 domains of cognitive functions, we found associations between CNVRs and fluid intelligence (B), working memory (C) and reading ability (D) after multiple testing correction (p<5×10^−4^ divided by 6, which is the effective number of tests across 8 dimensions of cognitive function). Dashed line indicates the threshold of genome-wide significance for each phenotype.

## Association of neurodevelopmental CNVs with psychopathology and cognition

To complement the genome-wide scan, we specifically looked for the presence of 48 deletions and 36 duplications that have previously been identified as recurrent CNVs associated with genetic risk for neurodevelopmental disorders (Supplementary Table 19 and Supplementary Figure 8)^22^. Among the 4111 individuals from the ABCD**^®^** study, we identified 20 deletions among 30 ABCD**^®^**individuals who showed at least 50% reciprocal overlap^58^ with 9 out of 48 neurodevelopmental deletions (Supplementary Table 19). 64 individuals carried 22 duplications showing at least 50% reciprocal overlap with 10 of 36 neurodevelopmental duplications (Supplementary Table 19). Of note, 59 out of 93 individuals carrying neurodevelopmental CNVs had either a deletion at 15q11.2, or a duplication at 15q11.2 or 16p13.11 (Supplementary Table 19). Next, we separately examined whether individuals carrying neurodevelopmental CNVs predicted each psychopathological and cognitive phenotype in the ABCD**^®^**study (see Methods). Our findings revealed that individuals with neurodevelopmental CNVs had high scores in the social domain of psychopathology (FDR-corrected p=1.55×10^−2^, Figure 4A, Supplementary Figure 9 and Supplementary Table 20). Moreover, these individuals showed poorer cognitive performance across various domains, including fluid intelligence (p_FDR_=1.47×10^−5^), attention (p_FDR_=1.75×10^−3^), working memory (p_FDR_=1.47×10^−5^), flexible thinking (p_FDR_=2.35×10^−4^), reading ability (p_FDR_=2.59×10^−2^) and processing speed (p_FDR_=2.65×10^−3^, Figure 4B, Supplementary Figure 9 and Supplementary Table 20).

**Figure 4.**
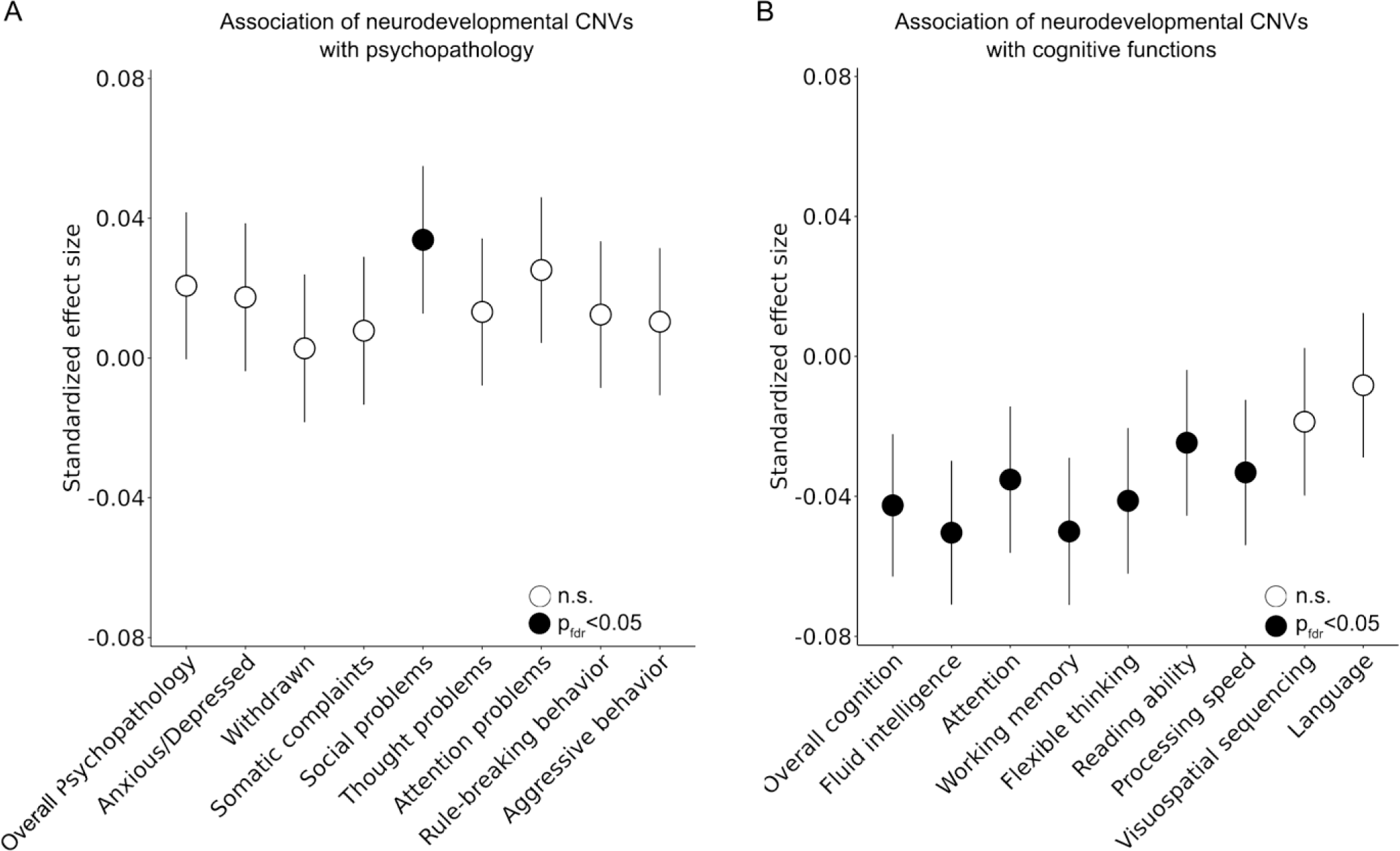
Associations of psychopathology and cognitive function with individuals carrying known pathogenic recurrent CNVs associated with neurodevelopmental disorders. (A) Association of overall and dimensional psychopathology with neurodevelopmental CNVs. Individuals with neurodevelopmental CNVs had significantly elevated psychopathology in the social domain at FDR-corrected p<0.05. (B) Association of global and dimensional cognitive function with neurodevelopmental CNVs. Individuals with neurodevelopmental CNVs had lower cognitive performance across domains including fluid intelligence, attention, working memory, flexible thinking, reading ability and processing speed (p_FDR_<0.05). Points in the dot plots indicate the value of a given predictor variable’s standardized effect size and error bars indicate 95%CIs for models of cognition and psychopathology.

### CNV risk score association with cognition and psychopathology

At the individual level, we quantified the burden of deletions based on five metrics: the size of deletions (mean=48.89kb [0-6.14Mb]), the number of genes covered (mean=0.13 [0-46]), pLI (mean=0.02 [0-12.77]), iLOEUF (mean=0.17 [0-77.89]) and pHI (mean=0.05 [0-24.27], see Methods and Supplementary Figure 10). For duplications, the CNV risk score at the participant level was determined by the size of duplications (mean=182.51Kb [0-11.10Mb]), the number of genes encompassed (mean=1.26 [0-51]), pLI (mean=0.11 [ranging 0-14.42]), iLOEUF (mean=1.23 [0-86.46]) and pTS (mean=0.295 [0-26.77]). Note that individuals who have no CNVs have CNV risk scores of 0.

To assess the cumulative effect of CNVs on psychopathology and cognition, regression models were performed with each CNV risk score across 8564 individuals, for global and dimensional psychopathology and cognition measures. For deletions, we observed that all domains of the CNV risk score were significantly associated with global cognition and all cognitive subdomains with the exception of language processing (p_FDR_<0.05, Figure 5 and Supplementary Tables 21 and 22; significant subdomains included fluid intelligence, attention, working memory, flexible thinking, reading ability, processing speed, visuospatial sequencing). Deletion CNV risk scores with the exception of CNV size were also significantly associated with overall psychopathology as well as the social, thought, and attention domains (p_FDR_<0.05, Figure 5 and Supplementary Tables 23 and 24). For duplications, iLOEUF and pLI risk scores were significantly associated with decreased overall cognition. Duplication risk scores were not significantly associated with domains of psychopathology after correction for multiple comparisons.

**Figure 5.**
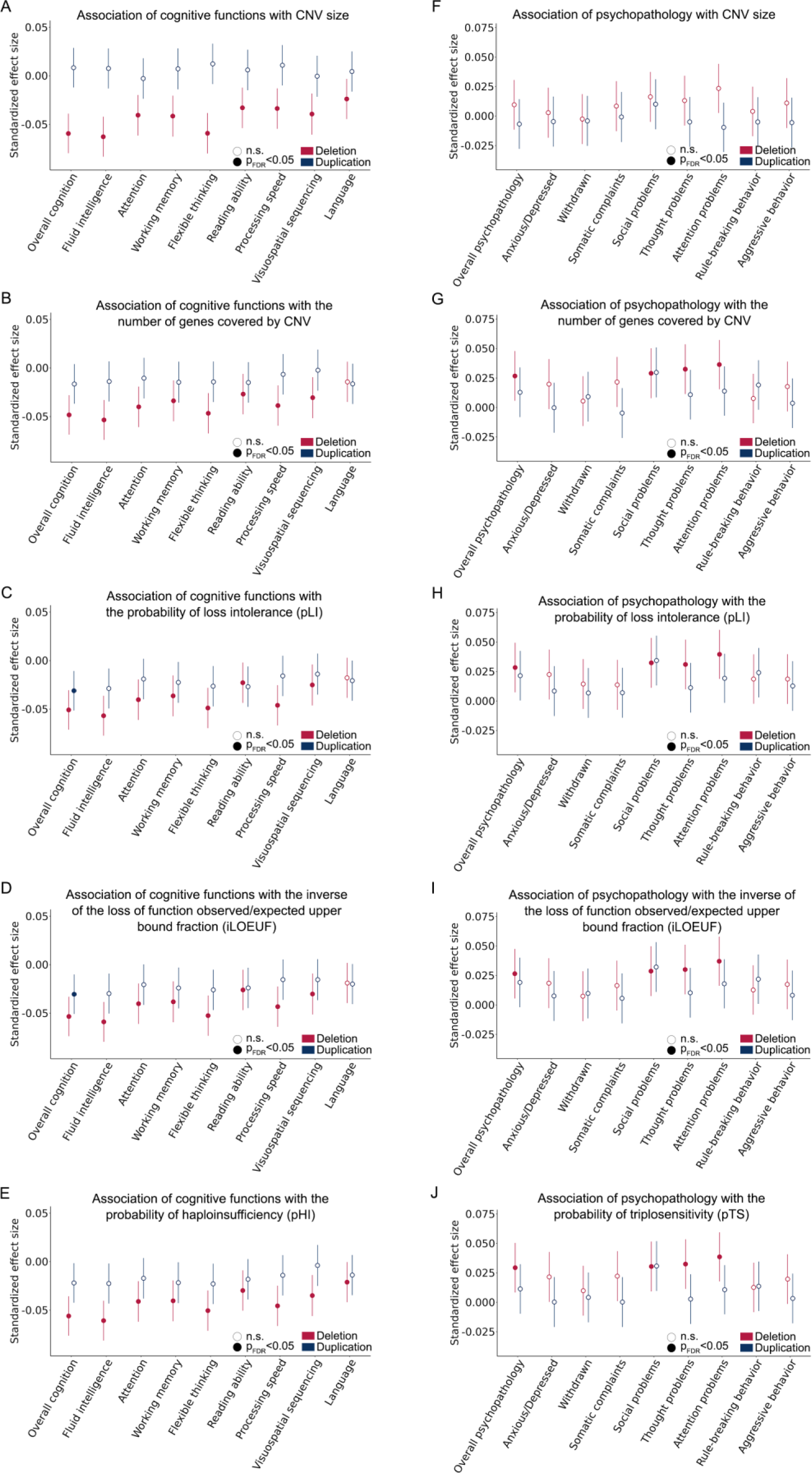
Associations of psychopathology and cognitive function with CNV risk scores. To determine the cumulative effect of CNVs on psychopathology and cognition, we used regression models to test associations with global and dimensional cognitive functions for each of 5 CNV risk scores across 8564 individuals, including the size of CNV (A), the number of genes overlapping with the CNV (B), the probability of loss intolerance (C), the inverse of the loss of function observed/expected upper bound fraction (D), dosage sensitivity (E). Similarly, we also tested associations with global and dimensions of psychopathology for each of the 5 CNV risk scores (F-J). Points in the dot plots indicate the value of a given predictor variable’s effect size and error bars indicate 95%CIs for models of cognition and psychopathology.

## Discussion

The present study delineates the landscape of CNVs in the ABCD**^®^**study and characterizes CNVRs associated with child psychopathology and cognitive function. Individuals in the ABCD**^®^**study carrying recurrent CNVs previously implicated in neurodevelopmental disorders exhibited greater psychopathology in the social domain and lower cognitive performance particularly in the domain of working memory. Aggregated CNV risk scores for deletions were associated with social, thought, and attention domains of psychopathology, as well as with nearly all cognitive domains. Additionally, duplication CNV risk scores derived from constraint scores (pLI and iLOEUF) were associated with lower overall cognitive function. These results underscore the significant impact of genomic structural changes on psychopathology and cognitive development in childhood. Furthermore, our study highlights the sensitivity of CNV risk scores as a measure to characterize the complex genetic underpinnings of neurodevelopmental phenotypes.

Previous genetic studies have traditionally focused on identifying genomic structural variants associated with the risk of categorical psychiatric disorders using case-control designs^59,60^. However, case-control designs may not adequately capture the complexity of psychiatric symptoms characterized by high levels of comorbidity and ambiguous diagnostic boundaries^19,21,61^. In our current study, we sought to characterize CNVRs associated with dimensional psychopathology and cognitive outcomes in childhood. This dimensional approach allowed us to explore how gene dosage imbalance affects dimensional psychopathology in community population that was not ascertained for mental health problems. As compared to pathogenetic CNVs discovered in case-control studies, CNVs with a higher frequency in the general population are more likely to be inherited, and may modulate psychopathology and cognitive development without necessarily leading to clinically diagnosed mental illness or intellectual disability.

The colocalization of SNP and CNV association signals revealed weak evidence of common genetic variants (from previous GWAS of psychiatric disorders and cognition) within CNVRs. In the context of previous evidence of enriched GWAS signal within pathogenic CNVs^31^, this finding has several potential explanations. First, it is possible that SNPs tag a genetic haplotype with an inherited CNV, such that the GWAS SNP effect is capturing the effect of this CNV. Second, common genetic polymorphisms with subtle effects, acting as genetic modifiers of complex phenotypes, could act cumulatively with CNVs and environmental factors to modulate variability in psychopathology and cognitive ability during childhood. Third, both CNVs and SNPs may influence phenotypes through regulatory effects on nonadjacent genomic regions, which complicates the interpretation of simple overlap between CNVRs and GWAS summary statistics. Fourth, the psychiatric GWAS data we referenced originated from a meta-analytic study pooling results across 12 psychiatric disorders defined by a traditional dichotomous diagnosis framework^49^. Consequently, the genetic liability for adult psychiatric disorders may not fully reflect the genetic architecture underlying subclinical psychiatric symptoms during adolescence. Similarly for the intelligence GWAS, the results were derived from previous genome-wide meta-analytic studies evaluating overall general intelligence in adults^50^. However, cognitive abilities undergo significant developmental changes during childhood and adolescence, characterized by the rapid growth of neural circuits underlying various cognitive processes^36,62^. Consequently, the genetic basis of cognitive function in childhood may diverge from that in adulthood, such that existing GWAS fail to fully capture the complexity and nuances of cognitive function during adulthood.

In our investigation of CNVRs associated with psychopathology measures derived from the Child Behavioral Checklist, we identified 2 CNVRs that met conservative genome-wide threshold of statistical significance. A copy number gain at 10q26.3 was associated with higher overall psychopathology, and a copy number gain at 14q11.2 was associated specifically with the attention domain. Of note the CNVR on chromosome 10 associated with overall psychopathology did not meet our threshold for statistical significance for any specific domain, although a marginal association was observed with the somatic domain. The fact that relatively few CNVRs were identified illustrates that ABCD**^®^**, despite being a large study by the standards of developmental neuroscience, is relatively underpowered to fully characterize the complex architecture of CNV associations. A related challenge is that the Child Behavioral Checklist, which is based on parent report of items on a 3 point scale, provides a relatively limited depth of phenotypic characterization^37^. More sensitive phenotypic characterization by aggregating measures across future ABCD**^®^** time points may yield increased power to identify CNVRs associated with developmental psychopathology.

In our investigation of CNVRs associated with cognitive measures, we identified 3 CNVRs that met genome-wide significance after correction for multiple comparisons, which partially overlapped with the CNVRs associated with psychopathology. The CNVR chr14:20247780-20423950 associated with the attention domain of psychopathology largely overlapped with chr14:20209121-20426704, which was associated with global cognition as well as fluid intelligence and reading domains (reciprocal overlap: 80.97%). Furthermore, the genes encompassed by both regions were fully overlapping. One plausible explanation for this observation is that the variant encompasses pleiotropic genes that influence cognition and psychopathology, which are known to be inter-related^63^. On chromosome 10, we found psychopathological and cognitive measures were significantly associated with duplications of two CNVRs at 10q26.3, which encompasses *PPP2R2D* and *SYCE1*. Genetic variants and modifications of *PPP2R2D* have been reported to impact psychopathology and neural development^47,48,64^, and SYCE1 variants have been associated with Alzheimer’s Disease^65^. In addition, we recovered a genome-wide significant association between lower general cognitive function and a deletion at chr1:146485371-147995694. Fully overlapping with a known genomic hotspot at 1q12.1, a susceptibility locus for neurodevelopmental disorders^55,66^, the recovery of this CNVR provides important validation for the ParseCNV2 methodology. Individuals with 1q12.1 deletion are known to exhibit a range of physical and mental disorders across multiple scales, including congenital heart abnormality^67^, ligamentous laxity or joint hypermobility^68^, intellectual disability^54^, microcephaly^55^, developmental delay^55^, and cognitive deficits^54^.

Extending our analysis to other known recurrent, pathogenic CNVs associated with neurodevelopmental disorders^22^, we found that individuals carrying neurodevelopmental CNVs exhibited lower performance across almost all cognitive domains, a finding that is consistent with recent adult studies^69^. However, with the exception of deficits in the social domain, individuals carrying neurodevelopmental CNVs did not have significant differences in measures of psychopathology (Figure 4). The lack of more consistent effects on psychopathology may be indicative of heterogeneity in psychiatric presentation of neurodevelopmental CNVs, as compared to relative homogeneity of the cognitive impact across domains. It is also possible that the cognitive impact is more apparent during childhood (or more readily measured with structured batteries). Some individuals with current cognitive deficits may eventually progress to clinically diagnosed psychiatric disorders, as additional psychiatric manifestations may unfold on a developmental timeline across adolescence and adulthood, depending on interactions with environmental and other genetic factors (e.g., risk due to common genetic variants).

Compared to identifying specific CNVRs or querying known recurrent CNVs, CNV risk scores offer a complementary approach based on the expected functional consequences aggregated across CNVs, particularly in terms of encompassed genes’ tolerance to loss-of-function mutations and dosage sensitivity^22,23^. In our analysis of the association between CNV risk scores and psychopathology, we observed that deletion risk scores were associated with variations in overall psychopathology as well as social, thought, and attention domains in particular. However, similar associations were not observed with any duplication risk scores, despite duplications at 10q26.3 and 14q11.2 being associated with psychiatric symptoms in the genome-wide CNVR association analysis. One potential explanation for this discrepancy is that the effect sizes of duplications is generally lower and also more heterogenous compared to the effect sizes of deletions^70,71^. Interestingly, unlike other components of CNV risk scores, we found that the size of deletions did not significantly correlate with any psychopathological dimensions. This underscores the potential for improved CNV risk scores, possibly attuned to specific phenotypic associations, to be explored in future investigations.

Several methodological limitations should be noted in the present study. First, our CNV association results were constrained by the limited sample size. Small samples are more susceptible to false discoveries in genetic analysis^72^, particularly with rare genetic variants that may occur only once in a small population. However, given our strict quality control and the fact that the CNVR with the smallest sample size recovered a known pathogenic CNV, false discovery due to low sample size is unlikely to be an issue for the present study. More relevantly, small samples may overlook genetic loci influenced by the nuanced variability of phenotypes among individuals^73^. It is essential to collect larger child cohorts to replicate the current genetic findings, explore additional genomic structural variants related to cross-diagnostic psychiatric symptoms, and investigate how genetic events interact with environmental factors to modulate mental and cognitive development. Second, a technical limitation concerns the genotype platform used for inferring CNVs. Although this approach is generally considered reliable, it lacks SNP probes for detecting CNVs in repeat-enriched regions due to the complex properties of genomic architecture^32,74^. Employing advanced whole-genome sequencing platforms could enhance the detection of high-fidelity CNVs^75,76^. However, genotype array platforms remain the most cost-efficient option for calling reliable CNVs at the current stage. Third, while ABCD**^®^** used epidemiological sample strategies, the sample is not truly “population representative” and is likely to skew towards higher-functioning children. Consequently, children with severely affected CNVs were less likely to participate in the ABCD**^®^** study, leading to a decrease in power to detect CNVRs in the present study (e.g., compared to a clinically-ascertained sample) and an underrepresentation of the contribution of CNVs to neurodevelopmental phenotypes.

Cumulatively, the present results represent a significant advance in our understanding of CNVs in the ABCD**^®^** study. In sharing detected CNVs and analysis code for use by other researchers, this resource will be valuable in future work aimed at understanding CNVs’ impact on interindividual variability in complex traits including neurocognitive development and child psychopathology.

## Acknowledgements

The research was funded by R01MH132934 and R01MH133843. Data used in the preparation of this article were obtained from the Adolescent Brain Cognitive Development^SM^ (ABCD**^®^**) Study (https://abcdstudy.org), held in the NIMH Data Archive (NDA). The ABCD Study® is supported by the National Institutes of Health and additional federal partners under award numbers U01DA041048, U01DA050989, U01DA051016, U01DA041022, U01DA051018, U01DA051037, U01DA050987, U01DA041174, U01DA041106, U01DA041117, U01DA041028, U01DA041134, U01DA050988, U01DA051039, U01DA041156, U01DA041025, U01DA041120, U01DA051038, U01DA041148, U01DA041093, U01DA041089, U24DA041123, U24DA041147. A full list of supporters is available at https://abcdstudy.org/federal-partners.html. A listing of participating sites and a complete listing of the study investigators can be found at https://abcdstudy.org/consortium_members/. ABCD**^®^** consortium investigators designed and implemented the study and/or provided data but did not necessarily participate in the analysis or writing of this report. This manuscript reflects the views of the authors and may not reflect the opinions or views of the NIH or ABCD consortium investigators.

## Conflict of interests

AFA-B receives consulting income from Octave Bioscience. AFA-B and JS hold equity in and serve on the board of Centile Bioscience.

## Data Availability

CNV calls will be made available on NDAR (https://nda.nih.gov/study.html?id=2589), and code to derive CNVs and perform quality control is publicly available on Github (https://github.com/BGDlab).

## Notes

### Author Declarations

The study used ONLY openly available human data that were originally located at https://abcdstudy.org/.

